# The PBL teaching method in Neurology Education in the Traditional Chinese Medicine undergraduate students: An Observational Study

**DOI:** 10.1101/2022.11.10.22282173

**Authors:** Yun Jin Kim

## Abstract

**Objective:** To study the effect of the Problem Based Learning method in Neurology education for Traditional Chinese Medicine (TCM) undergraduate students.

**Methods:** In this observational study was conducted 2020/02 and 2020/04 intake the year three TCM undergraduate students of the School of Traditional Chinese Medicine, Xiamen University Malaysia. A total of 86 were enrolled in this study. They were randomly divided into conventional learning groups and PBL groups. Students who missed more than one session of the course and those who did not complete the questionnaires in the evaluation periods were excluded from the study (n=0). An independent sample t-test was used to compare the results between the two groups. A p-value <0.05 was considered significant.

**Results:** The PBL group was significantly effective for the students’ theoretical and clinical practical examination scores, the satisfaction of teaching level, students’ perspectives, and self-learning skills, as well as significantly higher DREEM scores than students who participated with the conventional group (*p*<0.05).

**Conclusion:** The PBL teaching method in Neurology education for TCM undergraduate students can involve an interesting learning method, significantly improve their learning performance, and the ability to analyze the problem-solving skills in the neurology disease and its management knowledge.

## Introduction

Traditional Chinese Medicine (TCM) education is an important educational component of medical educational system in Malaysia. The Traditional and Complementary Medicine (T&CM) Council, Ministry of Health, and Ministry of Higher Education stipulates undergraduate TCM higher education providers should following the programme standards. According to the programme standards criteria for the TCM education include fundamental courses in TCM, western medicine, and clinical practice et al. unfortunately, there is a shortage of neurology education components in current TCM educational system in Malaysia.

Neurology is perceived as the most difficult specialty by medical and TCM students, also non-specialist healthcare professionals, throughout the medical programme and beyond, globally [1]. In this difficulty, students and healthcare professionals, often find neurology less interesting, especially when most neurological conditions are the first visit by general practitioners and other healthcare professionals [2]. It may make neurology disease information less appealing for medical and TCM students or general practitioners wishing to pursue neurology as a career. Promoting TCM student interest in neurology education is an important way to better provide neurology disease management [3].

Problem-based learning (PBL) has been widely used in medicine fields and educational context to promote critical thinking and problem-solving in authentic learning outcomes [4]. It is a pedagogical approach whereby issues are explained within a scenario to enable medical students to identify their own learning objectives [5], and involves students working in small groups through a specific problem or set of problems, with the oversight of a lecturer. The problem solving within the group generates topics for individual self-study, after that the group reconvenes to discuss the topic further [6]. Lecturers act as facilitators and guides, it is the lecturers does not provide knowledge or information directly. Lecturers are encourage skills of inquiry by appropriate probing problems or questions of the students and encouraging students to develop the knowledge and critical thinking pathway to ask themselves and their peers’ to learn new knowledge and information [7]. The PBL is recognized as a successful innovative learning method in undergraduate medical education system, also effective in enhancing students’ clinical practice performance [8], and analytical skills [9].

In this observational study, we conducted PBL learning method for the Neurology education in the TCM undergraduate students. We evaluated whether it can make up for the shortcomings of conventional teaching, improve students’ learning interesting and students’ basic knowledge of neurology disease management.

## Methods

### Study design and setting

The cross-sectional online survey was collected at Xiamen University Malaysia, Malaysia. In the 2022/04 academic semester, the course titled “Hello, Neurology” was the reformed from the conventional learning methods to a PBL methods, it is designed and approved from the external examiner, registered neurologist, and certified the PBL instructors. The School of Traditional Chinese Medicine confirmed the content validity of each course contents and carried out the interventions in total eight sessions, each sessions conducted two hours for each group. The general course objectives are introduce the neurology and disease management to TCM undergraduate students.

### Ethical considerations

This study was approved by the Research Ethics Committee of the Xiamen University Malaysia (no. REC-2208.01). All participants completed informed consent form that included the study purpose before start of the study. All participants were reassured of the confidentiality and the right to withdraw at any time during the study.

### Participants

The comparative study was conducted 2020/02 and 2020/04 intake the year three TCM undergraduate students of the School of Traditional Chinese Medicine, Xiamen University Malaysia. A total of 86 were enrolled in this study. They were randomly divided into conventional learning group and PBL group. Students who missed more than one session of the course and those who did not completed the questionnaires in the evaluation periods were excluded from the study (n=0).

All lecturers were trained and certified the PBL senior lecturers/Assistant Professor/Associate Professor with registered teaching permit holders in the Ministry of Higher Education Malaysia.

### Training of the PBL lecturers

Between November 2021 and February 2022, the School of Traditional Chinese Medicine, Xiamen University carried out two day introductory courses on PBL principles for 12 academic staffs to involved in curriculum development, and on willingness to learn and hands-on teaching practice the PBL approach. The training learning objectives were to introduction, application, development and setting the PBL approach and simulate teaching practice, and tutorial sessions. During the training, academic staffs to understand and demonstrated PBL context and curriculum development using the focusing small group discussion. At the same time discussed the learning materials and evaluation questionnaire tools.

### Conventional and PBL teaching

We chose neurology disease and management contents as the topic for applying the conventional and PBL teaching approach in this study.

The faculty was conducted the conventional group was arranged as following:

The course lecturers prepared lecture slides and supplementary materials provide the students. The student were received about the disease definition, epidemiology, pathogenesis, differential diagnosis, disease management and prevention was carried out. It is course lecturer provided a thorough explanation of the theoretical medical knowledge, and the students answered the questions and made suggestions.

The PBL groups was arranged as following:

Firstly, the course lecturers conducted the disease definition and disease management and put forward to representative and enlightening questions according to the characteristics of the neurologic disorders.

Secondly, during the small group discussions, the students under the PBL lecturers’ monitoring and guidance. The students were encouraged to raise relevant clinical questions and problems, and searched the answers on the Internet, consult main reference books, and connect the university library database or other relevant materials. This process has been described as the seven classical steps of PBL. (1) Knowledgeable and understand the situation and clarify terminology; (2) Identify the questions and problems; (3) Suggest possible causes; (4) Connect questions and problems and causes; (5) Decide what type of information is needed; (6) Obtain relevant information; (7) Apply the information. Depending on the complexity of the questions and problems, additional mentoring may be required as the group narrows the possible solutions [10].

Thirdly, the student representative from each group to demonstrate to review the main points from the lessons, posed their group’s problems and questions and its relevant solving, and Q&A about unsolved problems and questions.

Finally, the PBL lecturers summarized the class and went over the problems and questions.

### Outcome measurement

All participants received theoretical and clinical practical skill examinations before and after completed learning. The theoretical written examination contents include the disease definitions, clinical symptoms, treatment and prevention of neurology disorders. The clinical practical skill examination incudes the diagnosis and differential diagnosis of neurology disorders, selection of treatment procedures. Both examination are 10 single choice questions with 1 point for each question.

Focusing on the investigation of satisfaction with teaching methods, the teaching method effect was evaluated through a feedback survey. The satisfaction with the teaching method included high-level satisfaction (above 80 points), general satisfaction (60 – 80 points), and dissatisfaction (less than 60 points). Then, the satisfaction degree was calculated according to the formula [11]: satisfaction degree = satisfaction rate + general rate.

The anonymous survey concerning the teaching methods questionnaires was conducted among all participants end of the course. The survey questionnaires including the students’ perspectives and self-perceived competence in the PBL and conventional groups and found that the scores for knowledge and understanding, Cognitive, lecturer-student interaction, communication skills, clinical practical skills, self-learning skills, teamwork skills, leadership skills, and ethics and professionalism. All questionnaires was adopted previous articles [12, 13]. All answers were based on a 5-point Likert scale, and 1 represented the least and 5 the highest degree of agreement.

The students’ perceptions of learning environment survey concerning the teaching methods by PBL and conventional teaching were sought by the use of a survey questionnaires was adopted Dundee Ready Educational Environment Measure (DREEM), it was designed to measure the educational environment specifically for medicine and other health professionals [14]. There were fifty items in the questionnaire to assess for the students’ perceptions of learning environment. Each relates to one of five themes: (1) Students’ perceptions of learning (12 items); (2) Students’ perception of teachers (11 items); (3) Students’ academic self-perceptions (8 items); (4) Students’ perceptions of the learning atmosphere (1 items); (5) Students’ social self-perceptions (7 items). Responses to the items are scored from 4 to 0, and 0 represented strongly disagree and 4 represented strongly agree. However, reverse scoreing was required for items 4, 8, 9, 17, 25, 35, 39, 48, and 50. The DREEM score is calculated between 0 to maximum score of 200 [15, 16]. According to for Medical Education in Europe (AMEE) ‘s guideline interpreted, a score of 51 – 100 indicates “plenty of problems”, while a score of 101 to 150 is “more positive than negative” and score of 100 is interpreted as “considerable ambivalence by students and needs to be improve”. Items with mean scores of less than two should be closely considered. Except for negative items mentioned, a higher score means better interpretation [17, 18].

### Statistical methods

The data collection and statistical analysed using the SPSS software version 16.0 for Windows (IBM, USA). All data are shown as mean ± standard deviation (SD). Applying the independent sample *t*-test was used to compare results between the PBL and conventional groups. Statistical significance was inferred at *p* values < 0.05.

## Result

Eighty-six participants were present at the initial session, and all eighty-six entered the study and were equally allocated into the conventional and PBL groups, including forty-three participants in each group. All eighty-six participants completed the theoretical and clinical practical examinations and questionnaires. Figure 1 shown the CONSORT study flow diagrams for this study.

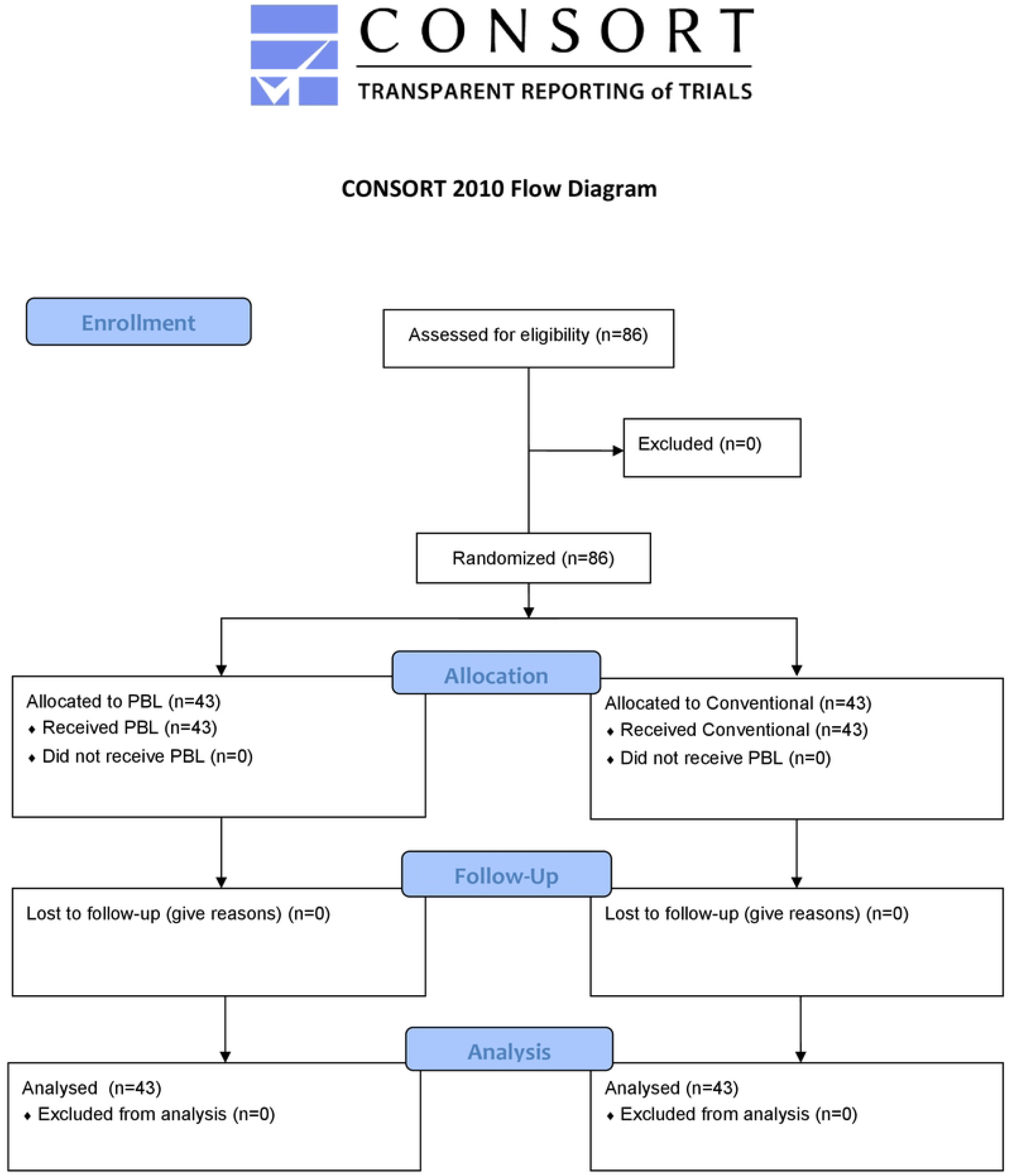

### Participants’ socio-demographics

A total 86 participants were enrolled in this study. They were randomly divided into PBL group (n=43) and conventional group (n=43). There were 20 (46.5%) male and 23(53.5%) female participants in the PBL group, in conventional group were 24 (55.8%) male and 19(44.2%) female participants. There are 10(23.2%) in the PBL and 8(18.6%) in the conventional groups who were scholarship holders. Total 36(83.7%) Malaysian and 7(16.3%) Chinese in the PBL group, There are 38(88.3%) Malaysian and 5(11.7%) Chinese in the conventional group. The average age of PBL group were 22.7±2.1 and conventional group were 22.6±1.8 years respectively. There was no significant difference in age, gender, scholarship, and nationality (*p*>0.05, data shown the table 1.

**Table 1.** Socio-demographic characteristics of the participants in the study (n=86).

|  |  | **PBL**<br>**(n=43)** | **Conventional**<br>**(n=43)** | ***p*-value** |
| --- | --- | --- | --- | --- |
| **Gender** |  |  |  | 0.716 |
|  | Male | 20 (46.5%) | 24 (55.8%) |  |
|  | Female | 23 (53.5%) | 19 (44.2%) |  |
| **Scholarship** |  |  |  | 0.629 |
|  | Yes | 10 (23.2%) | 8 (18.6%) |  |
|  | No | 33 (76.8%) | 35 (81.4%) |  |
| **Nationality** |  |  |  | 0.896 |
|  | Malaysian | 36 (83.7%) | 38 (88.3%) |  |
|  | Chinese | 7 (16.3%) | 5 (11.7%) |  |
| **Age (Year, Mean ± SD)** |  | 22.7±2.1 | 22.6±1.8 | 0.901 |

### The comparison of theoretical and clinical practice examination scores between the PBL and conventional groups

The Table 2 shown the before and after Theoretical and Clinical practical skill examination scores between the PBL and conventional groups. In the conventional group, the before Theoretical examination score were 3.39±1.51 and Clinical practical skill examination score were 4.05±1.56; after Theoretical examination score were 6.01±1.46 and Clinical practical skill examination score were 6.98±1.23. In the PBL group, the before Theoretical examination score were 3.27±1.65 and Clinical practical skill examination score were 3.86±1.83; after Theoretical examination score were 8.35±1.39 and Clinical practical skill examination score were 7.79±1.52 respectively. There are no statistical significance in pre Theoretical examination and Clinical practical skill examination scores both groups (*p*>0.05). The after Theoretical examination and Clinical practical skill examination scores in the PBL group were significantly higher than conventional group (*p*<0.05).

**Table 2.** Comparison of the Theoretical and Clinical practice examination scores between the PBL and conventional groups.

|  | PBL<br>(n=43) | Conventional<br>(n=43) | *p*-value |
| --- | --- | --- | --- |
| **Before** |  |  |  |
| Theoretical examination | 3.27±1.65 | 3.39±1.51 | 0.162 |
| Clinical practical skill examination | 3.86±1.83 | 4.05±1.56 | 0.396 |
| **After** |  |  |  |
| Theoretical examination | 8.35±1.39* | 6.01±1.46 | 0.019 |
| Clinical practical skill examination | 7.79±1.52* | 6.98±1.23 | 0.024 |

Values represent mean ± SD.
\* Indicates a statistical difference from the conventional group (*p*<0.05).

### The satisfaction teaching level of TCM students with different teaching groups

Comparing the satisfaction teaching level of TCM students in the PBL and conventional groups, we found the overall satisfaction teaching level of the PBL group was significantly higher than conventional group (*p*<0.05). The data shown in the Table 3.

**Table 3.** The satisfaction teaching level of TCM students with different teaching methods

|  | Satisfaction | General | Dissatisfaction | Satisfaction Degree | *p*-value |
| --- | --- | --- | --- | --- | --- |
| **PBL**<br>**(n=43)** | 38 | 3 | 2 | 95%* | 0.038 |
| **Conventional**<br>**(n=43)** | 18 | 16 | 9 | 79% |  |

\* Indicates a significant difference from the conventional group (*p*<0.05)

### The questionnaire survey scores between the PBL and conventional groups

We conducted after course survey scores concerning to students’ perspectives and self-learning competence in the PBL and conventional groups. We found the cognitive, lecturer-student interaction, communication skills, clinical practical skills, self-learning skills, teamwork skills, and leadership skills were significantly higher in the PBL group than in the conventional group. The results shown in the Table 4.

**Table 4.** Questionnaire survey scores between the PBL and Conventional groups.

|  | PBL<br>(n=43) | Conventional<br>(n=43) | *p*-value |
| --- | --- | --- | --- |
| Knowledge and Understanding | 5.12±0.87 | 5.01±0.56 | 0.062 |
| Cognitive | 4.02±0.28* | 3.03±0.17 | 0.015 |
| Lecturer-Student interaction | 4.99±0.82* | 2.94±0.74 | 0.024 |
| Communication skills | 4.89±0.76* | 2.85±0.81 | 0.014 |
| Clinical Practical skills | 4.78±0.68* | 2.57±0.81 | 0.012 |
| Self-learning skills | 4.07±0.79* | 2.42±0.63 | 0.017 |
| Teamwork skills | 3.96±0.82* | 2.40±0.81 | 0.031 |
| Leadership skills | 4.08±0.51* | 2.51±0.65 | 0.029 |
| Ethics and Professionalism | 4.13±0.91 | 4.08±0.82 | 0.071 |

Values represent mean ± SD.
\* Indicates a statistical difference from the conventional group (*p*<0.05)

**Table 5.** The comparison of DREEM scores between the PBL and conventional groups

| **DREEM** | **Max. Score** | **PBL**<br>**(n=43)** | **Conventional**<br>**(n=43)** | ***p*-value** |
| --- | --- | --- | --- | --- |
| **SPL** | 48 | 31.37±7.1* | 27.39±7.3 | 0.012 |
| **SPT** | 44 | 27.07±5.9* | 24.78±5.6 | 0.017 |
| **SAP** | 43 | 20.44±5.5* | 18.68±5.1 | 0.015 |
| **SPA** | 48 | 27.72±8.8 | 27.47±6.8 | 0.063 |
| **SSP** | 28 | 15.88±4.9* | 10.55±4.1 | 0.023 |
| **Total DREEM score** | 200 | 122.58±25.6* | 115.91±22.7 | 0.019 |

Values represent mean ± SD.
\* Indicates a statistical difference from the conventional group (*p*<0.05)
Abbreviations: DREEM, Dundee Ready Education Environment; SPL, Students’ Perception of Learning; SPT, Students’ Perception of Teachers; SAP, Students’ Academic Self-Perception; SPA, Students’ Perception of Atmosphere; SSP, Students’ Social Self-Perception.

### The comparison of DREEM scores between the PBL and conventional groups

The total DREEM scores for the PBL group was significantly higher than the conventional group. We found the Students’ Perception of Learning, Students’ Perception of Teachers, Students’ Academic Self-Perception, and Students’ Social Self-Perception scores were significantly higher in the PBL group than the conventional group. However, Students’ Perception of Atmosphere score were not significantly difference between the PBL and conventional groups.

## Discussion

Neurology is a branch of general medicine dealing with disorders of the nervous system, is considered one of the most intimidating and complicated fields of the medical sciences by medical students and general practitioners [19]. Understanding and being well-trained in neurology could spend considerable time and effort, in fact, it is a lack of systematic and comprehensive neurology education for undergraduate students and continuing education programme for general practitioners or other related healthcare professionals, who wrongly understand neurology disorders and management [20].

TCM education in Malaysia is divided into two concepts – academic and skill-based education. The main educational aim of this is the quality control of higher education providers that provide high-impact academic education. As the higher education provider of TCM to overcome the challenges associated with the need to improve the resources required to ensure the quality of high-impact education [21, 22]. Moreover, the TCM students stipulate stronger integrative medical concepts to completing the regular curriculum. Although this provision has not yet been implemented, such strong integrative medical concepts also advanced the level of education for the new generation of TCM practitioners who are knowledgeable in biomedical sciences, western medical sciences, and well-versed in TCM and at the same time adequately educated medical law, medical ethics, professionalism, and humanities [23].

PBL education starting from learning is a collaborative process anchored in student groups. The principles further state how medical students are responsible for their own personal learning while being supported by one or more lecturers. It is also emphasized that problems must be exemplary and scientific. Problems must therefore reflect conditions realistic and authentic within a medical academic field or relevant to a profession [24]. Therefore, PBL education is concerned with the future professions of medical students and promotes metacognitive skills by having medical students engage with authentic and complex problems. Thus, PBL is reflected in competencies such as the ability to self-learning, to collaborate with group members, and to initiate and organize discussion when encountering complex real-life problems during clinical practice [25].

This observational study compared the PBL and conventional learning methods for neurology education in TCM undergraduate students using theoretical and clinical practical examination scores, the satisfaction with teaching level, students’ perspectives and self-learning skills, and the Dundee Ready Education Environment Measure (DREEM). When the PBL group was significantly effective for the students’ theoretical and clinical practical examination scores, the satisfaction of teaching level, students’ perspectives, and self-learning skills, as well as significantly higher DREEM scores than students who participated with the conventional group.

## Conclusion

On the basis of the results of the present study, it can be concluded that the PBL teaching method is preferable for neurology education in TCM undergraduate students over the conventional teaching method. The PBL teaching method can support and stimulate the learning interest of TCM undergraduate students, and improve the ability of logical thinking and problem-solving skills and solutions. Therefore, this study highlighted the PBL teaching method is a highly valued educational format and has been broadly applied in neurology education, such as for the retention and application of neurology disease and its clinical management knowledge. The limitation of the current study can’t conduct double-blind research and compared other traditional medical education teaching methods, such as case-based learning methods. Also needed to uncover the mechanisms by which the PBL learning method is more effective in neurology education in TCM undergraduate students. Such information should need to improve the PBL skills of the course lecturers, and explicitly develop neurology educational framework and skills to optimize learning outcomes.

## Data Availability

All relevant data are within the manuscript and its Supporting Information files.

## Acknowledgement

The authors thanks to all the TCM undergraduate students who volunteered to participate in the study, and Prof. Xin Gang, Department of Microbiology and Immunology, Shantou University, China to conduct the PBL training.

## Availability of data and materials

All data generated or analysed during this review are included in this article.

## Declarations

## Ethics approval and consent to participate

The ethics committee of the Xiamen University Malaysia approved this study. All methods were carried out in accordance with the institutional guidelines and regulations, and written informed consent was obtained from all participants.

## Consent for publication

Not applicable.

## Reference

[1] Pakpoor, J., Handel, A. E., Disanto, G., Davenport, R. J., Giovannoni, G., Ramagopalan, S. V., & Association of British Neurologists (2014). National survey of UK medical students on the perception of neurology. BMC medical education, 14, 225. https://doi.org/10.1186/1472-6920-14-225

[2] Kamour, A. H., Han, D. Y., Mannino, D. M., Hessler, A. B., & Kedar, S. (2016). Factors that impact medical student and house-staff career interest in brain related specialties. Journal of the neurological sciences, 369, 312–317. https://doi.org/10.1016/j.jns.2016.08.046

[3] Collin S., Khadijah M., Mark P. G., Shilpa C., & Hina Dave (2022). Education Research: Enhancing Medical Student Interest in Careers in the Clinical Neurosciences Through a Hands-on Procedure Workshop. Neurol Edu, 1 (1), e200010. http://doi.org/10.1212/NE9.0000000000200010

[4] Prosser, M., & Sze, D. (2014). Problem-based learning: student learning experiences and outcomes. Clinical linguistics & phonetics, 28(1-2), 131–142. https://doi.org/10.3109/02699206.2013.820351

[5] Bodagh, N., Bloomfield, J., Birch, P., & Ricketts, W. (2017). Problem-based learning: a review. British journal of hospital medicine (London, England : 2005), 78(11), C167–C170. https://doi.org/10.12968/hmed.2017.78.11.C167

[6] Jin, J., & Bridges, S. M. (2014). Educational technologies in problem-based learning in health sciences education: a systematic review. Journal of medical Internet research, 16(12), e251. https://doi.org/10.2196/jmir.3240

[7] Santos, M., Otani, M., Tonhom, S., & Marin, M. (2019). Degree in Nursing: education through problem-based learning. Revista brasileira de enfermagem, 72(4), 1071–1077. https://doi.org/10.1590/0034-7167-2018-0298

[8] Al-Azri, H., & Ratnapalan, S. (2014). Problem-based learning in continuing medical education: review of randomized controlled trials. Canadian family physician Medecin de famille canadien, 60(2), 157–165.

[9] Engel, C. E., Browne, E., Nyarango, P., Akor, S., Khwaja, A., Karim, A. A., & Towle, A. (1992). Problem-based learning in distance education: a first exploration in continuing medical education. Medical education, 26(5), 389–401. https://doi.org/10.1111/j.1365-2923.1992.tb00192.x

[10] Neve, H., Bull, S., Lloyd, H., Gilbert, K., & Mattick, K. (2018). Evaluation of an innovative, evidence-guided, PBL approach. The clinical teacher, 15(2), 156–162. https://doi.org/10.1111/tct.12656

[11] van der Vleuten, C., & Schuwirth, L. (2019). Assessment in the context of problem-based learning. Advances in health sciences education : theory and practice, 24(5), 903–914. https://doi.org/10.1007/s10459-019-09909-1

[12] Esan, T. A., & Oziegbe, E. O. (2015). Comparative evaluation of perceptions of dental students to three methods of teaching in Ile-Ife, Nigeria. African journal of medicine and medical sciences, 44(4), 335–341.

[13] Surratt, C. K., & Desselle, S. P. (2007). Pharmacy students’ perceptions of a teaching evaluation process. American journal of pharmaceutical education, 71(1), 6. https://doi.org/10.5688/aj710106

[14] Miles, S., Swift, L., & Leinster, S. J. (2012). The Dundee Ready Education Environment Measure (DREEM): a review of its adoption and use. Medical teacher, 34(9), e620–e634. https://doi.org/10.3109/0142159X.2012.668625

[15] Chan, C., Sum, M. Y., Tan, G., Tor, P. C., & Sim, K. (2018). Adoption and correlates of the Dundee Ready Educational Environment Measure (DREEM) in the evaluation of undergraduate learning environments - a systematic review. Medical teacher, 40(12), 1240–1247. https://doi.org/10.1080/0142159X.2018.1426842

[16] Khalaf, H., Almothafar, B., & Alhalabi, N. (2022). Iraqi Medical Student’s Perceptions of Learning Environment Following Surgical Curriculum Change. Acta informatica medica, 30(2), 105–109. https://doi.org/10.5455/aim.2022.30.105-109

[17] Behkam, S., Tavallaei, A., Maghbouli, N., Mafinejad, M. K., & Ali, J. H. (2022). Students’ perception of educational environment based on Dundee Ready Education Environment Measure and the role of peer mentoring: a cross-sectional study. BMC medical education, 22(1), 176. https://doi.org/10.1186/s12909-022-03219-8

[18] Pelzer, J. M., Hodgson, J. L., & Werre, S. R. (2014). Veterinary students’ perceptions of their learning environment as measured by the Dundee Ready Education Environment Measure. BMC research notes, 7, 170. https://doi.org/10.1186/1756-0500-7-170

[19] Dong, L., Gao, T., Zheng, W., Zeng, K. & Wu, X. (2021), E-Learning for Continuing Medical Education of Neurology Residents. Mind, Brain, and Education, 15: 48–53. https://doi.org/10.1111/mbe.12271

[20] Mia, M., Sangida A., Mariana E-P., & Raddy R. (2022), Education Research: Bridging the Undergraduate Neurosciences With Clinical Neurology. Neuroscience Faculty Perspectives. Neurology Education, 1(1): 3200005. https://doi.org/10.1212/NE9.0000000000200005.

[21] Park, Y. L., & Canaway, R. (2019). Integrating Traditional and Complementary Medicine with National Healthcare Systems for Universal Health Coverage in Asia and the Western Pacific. Health systems and reform, 5(1), 24–31. https://doi.org/10.1080/23288604.2018.1539058

[22] Park, J. E., Yi, J., & Kwon, O. (2022). Twenty years of traditional and complementary medicine regulation and its impact in Malaysia: achievements and policy lessons. BMC health services research, 22(1), 102. https://doi.org/10.1186/s12913-022-07497-2

[23] Du H. B. (2015). Traditional Chinese medicine education in Canada. Chinese journal of integrative medicine, 21(3), 173–175. https://doi.org/10.1007/s11655-014-1963-7

[24] Stentoft D. (2019). Problem-based projects in medical education: extending PBL practices and broadening learning perspectives. Advances in health sciences education : theory and practice, 24(5), 959–969. https://doi.org/10.1007/s10459-019-09917-1

[25] Dolmans D. (2019). How theory and design-based research can mature PBL practice and research. Advances in health sciences education: theory and practice, 24(5), 879–891. https://doi.org/10.1007/s10459-019-09940-2

